# Current Limitations of Electronic Health Record Systems in Supporting Pragmatic Clinical Trials: Insights from the eMERGE Consortium

**DOI:** 10.1101/2025.04.01.25325049

**Authors:** Kavishwar B. Wagholikar, Jennifer Allen Pacheco, Adam S. Gordon, Atlas Khan, Bahram Namjou Khales, Barbara Benoit, Benjamin J. Kerman, Chunhua Weng, Casey Ta, Cynthia A. Prows, Robert Johnson, Dan M. Roden, David Crosslin, Elizabeth M. McNally, Elizabeth W. Karlson, Frank Mentch, Gail P. Jarvik, Georgia L. Wiesner, Hakon Hakonarson, James J. Cimino, Jeritt G.Thayer, Jordan W. Smoller, Jodell E. Linder, John Connolly, Josh F. Peterson, Josh Cortopassi, Krzysztof Kiryluk, Marwan Hamed, Mary Maradik, Megan J. Puckelwartz, Mohammadreza Naderian, Nephi Walton, Nita Limdi, Devi Priyanka Maripuri, Theresa Walunas, Vivian Gainer, Yuan Luo, Cong Liu, Eimear E. Kenny, Angelica Espinoza, Robb Rowley, Wei-Qi Wei, Shawn N. Murphy

**Author notes:** **Corresponding Author** Kavishwar B. Wagholikar, MBBS, PhD, 399 Revolution drive Somerville MA 02145.

## Abstract

Pragmatic clinical trials (PCTs) evaluate interventions in real-world settings, often using electronic health records (EHRs) for efficient data collection. We report on the challenges in performing EHR analysis of health-care provider orders in a PCT within the eMERGE consortium, which investigates the impact of reporting genome-informed risk assessments (GIRA) to over 25,000 patients across 10 academic medical centers. Clinical informaticians conducted a landscape analysis to identify approaches for evaluating the outcomes of GIRA reporting through the EHR. Of 98 identified outcomes, 54 (55.1%) were determined to be difficult to extract because they involved provider orders, which are typically documented in free text or proprietary formats within the EHR and only mapped to standardized codes after the service is completed. These findings highlight a critical barrier in using EHRs to support PCTs. The authors recommend closer collaboration between clinicians and informaticians, improved EHR systems that support standardized order entry, and future use of machine learning to automate analysis of provider behavior in clinical trials.

## Introduction

Pragmatic clinical trials (PCTs) are designed to evaluate the effectiveness of interventions in real-world practice by focusing on diverse populations, flexible protocols, and practical outcomes. [1] Unlike traditional trials in controlled environments, PCTs often leverage electronic health records (EHR) for cost-efficient enrollment and data collection. Here, we report on the challenges in using health-care provider orders in a PCT within the eMERGE consortium, which investigates the impact of reporting genome-based scores. [2] Analyzing provider orders is crucial to distinguish whether gaps in clinical care stem from provider inaction, or other barriers that preclude patients from following up on recommended care.[3]

## Methods

The study includes 25,003 patients recruited from medical practices across 10 academic medical centers. The participants provided electronic consent and age-based assent, completed surveys and submitted biological samples. These data were used to generate a “genome-informed risk assessment” (GIRA) report for 11 common diseases, which was shared with the patients and their healthcare providers. Clinical experts created disease-specific follow-up recommendations in the GIRA report that was returned to high-risk patients. Clinical experts also created a corresponding list of outcome variables for EHR extraction to determine the clinical impact of the GIRA report – whether the proportions of providers recommending risk-reducing interventions and patients undertaking interventions are higher amongst the high-risk patients. Clinical informaticians performed a landscape analysis to identify methods for extracting the outcome variables from the EHR.

## Results

The clinicians identified 98 outcomes for extraction from the EHR. The informaticians considered the use of the Observational Medical Outcomes Partnership (OMOP) Common Data Model (CDM) and terminology mappings for extracting EHR data from each site including demographic, visits, diagnoses, procedures, laboratory-test results and medications.[4] However, the informaticians determined that extracting 54 (55.1%) of the outcomes would be challenging, as they related to provider actions, such as ordering laboratory tests, procedures or referrals (see supplementary file). This difficulty arises because provider orders in EHR systems are recorded in the clinical data model but are not initially coded (see Figure 1).[5] Instead, they are documented using proprietary codes or free text during ordering and are mapped to Current Procedural Terminology (CPT) codes only after the ordered service is completed. Consequently, the group decided to extract the text descriptions associated with the orders and to manually map them to the outcomes.

**Figure 1.**
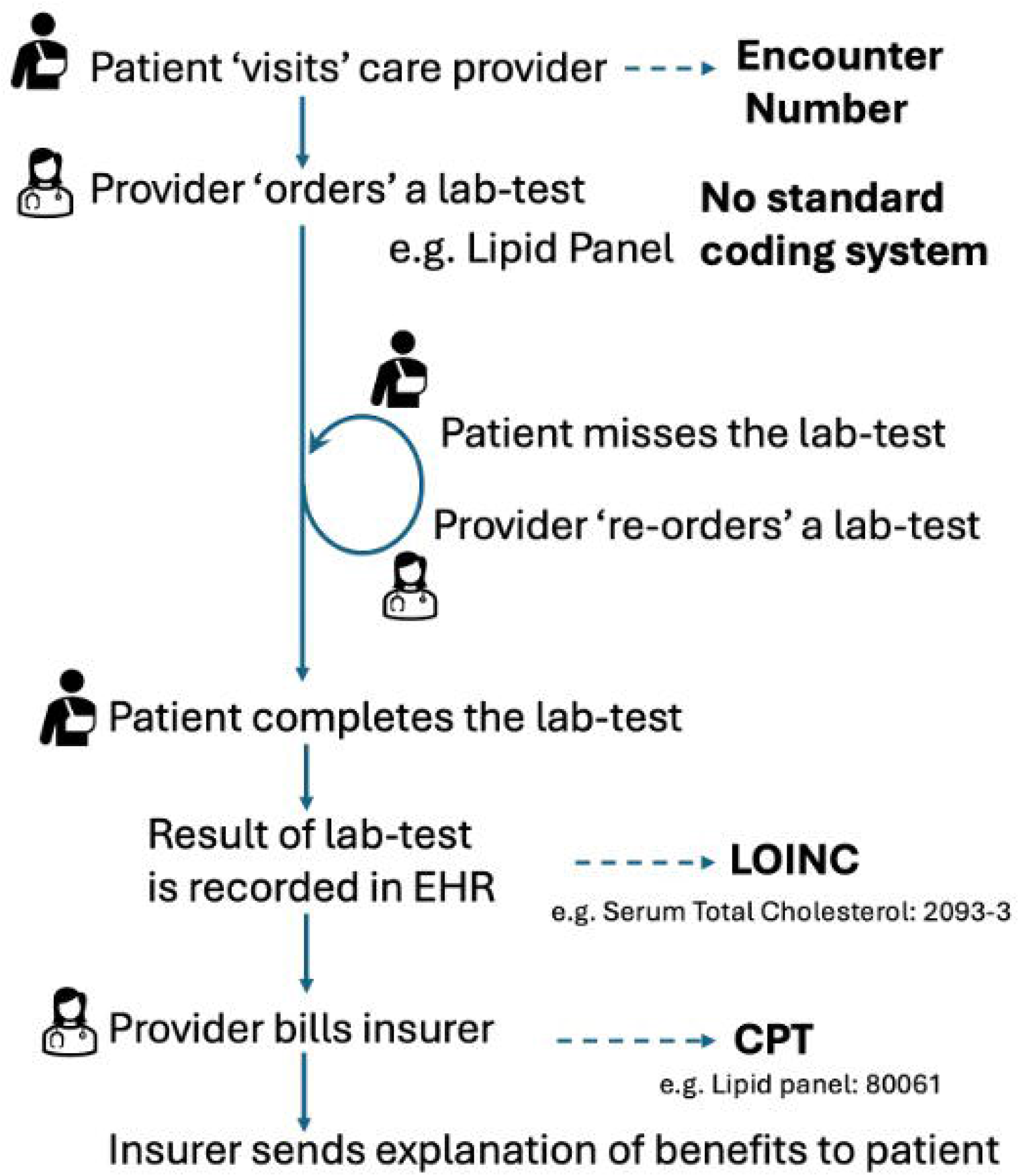
Provider orders for labs and procedures are recorded in the EHR, but they are not coded i.e., mapped to a standard terminology like CPT until after the procedure is completed or the results are reported.

## Discussion

Our study reveals that EHR systems are not inherently designed to facilitate the analysis of provider ordering, requiring extensive manual effort to determine if the provider adhered to the recommended actions. This finding highlights a major obstacle for pragmatic clinical trials, where it is often essential to decompose the clinical effects at both provider and patient levels– i.e. provider adherence to the study protocol or clinical guideline, and patient adherence to the provider recommendations. Patient behavior can be influenced by various factors, including disagreement with the provider, seeking follow-up care in a different healthcare system, and social determinants of health. Since many orders remain ‘not completed’ due to patient and system-related factors, relying solely on CPT code analysis may underestimate the true clinical impact of the intervention being evaluated in PCTs. To address this issue, we advocate for close collaboration between informaticians and clinicians to decipher the record of provider orders. Additionally, EHR vendors should integrate order entry systems with terminology databases or implement drop-down menus, for recording orders using standardized codes. Until such systems are implemented researchers will need to resort to manual methods to analyze the provider actions. Eventually the use of large language models [6] and machine learning may be useful to monitor and analyze the provider actions.

**Supplementary file 1**. The clinical outcomes identified by the clinical experts are associated with concepts that need to be extracted from the EHR to assess the outcomes. Among the 98 distinct outcomes, 54 (55.1%) are considered challenging to extract, as they require extracting data on provider actions, such as ordering laboratory tests, procedures, or referrals (as noted in Column F).

## Data Availability

All data produced in the present work are contained in the manuscript

## Acknowledgements

This phase of the eMERGE Network was initiated and funded by the NHGRI through the following grants: U01HG011172 (Cincinnati Children’s Hospital Medical Center); U01HG011175 (Children’s Hospital of Philadelphia); U01HG008680 (Columbia University); U01HG011176 (Icahn School of Medicine at Mount Sinai); U01HG008685 (Mass General Brigham); U01HG006379 (Mayo Clinic); U01HG011169 (Northwestern University); U01HG011167 (University of Alabama at Birmingham); U01HG008657 (University of Washington); U01HG011181 (Vanderbilt University Medical Center); U01HG011166 (Vanderbilt University Medical Center serving as the Coordinating Center). Dr. Wagholikar’s efforts were partially supported by R01-HL151643.

